# Change in Resting-State functional MRI Connectivity to Measure Individual Response to Epilepsy Surgery

**DOI:** 10.1101/2024.12.19.24319178

**Authors:** Emilio G. Cediel, Erika A. Duran, Jeffrey Laux, Olivia Leggio, William Reuther, Belfin Robinson, Angela Wabulya, Varina L. Boerwinkle

## Abstract

**Objective:** This study evaluates fractional power spectrum contribution (fPSC), a biomarker derived from resting-state functional MRI (rs-fMRI), as an indicator of epileptogenic network activity in drug-resistant epilepsy (DRE) patients undergoing surgery. We aimed to assess pre-to post-operative changes in fPSC and their correlation with seizure outcomes.

**Methods:** A retrospective cohort of 56 pediatric DRE patients with pre- and post-operative rs-fMRI were evaluated. Independent component analysis (ICA) was applied to identify resting-state networks (RSNs). Each ICA RSN’s power spectrum within the range of 0.06–0.25 Hz was quantified by fPSC. The change in this fPSC was compared pre- and post-operatively using paired t-tests. Multivariate analyses including correlations with clinical outcomes were evaluated by linear mixed effects models and ANOVA.

**Results:** Among the 56 patients, 80.4% demonstrated greater than 50% seizure reduction post-surgery, with 64.3% achieving seizure freedom. fPSC significantly decreased after surgery (t=3.0, p=0.005), indicating a reduction in epileptogenic network activity. The mixed effects model, controlling for covariates, also showed a significant effect of post-surgical scan on fPSC reduction (χ^²^=8.4, df=1, p=0.004). However, there was insufficient evidence to establish an association between changes in fPSC and clinical improvement score (p=0.16) or seizure frequency (p=0.49).

**Conclusion:** The observed reduction in fPSC post-surgery highlights its potential as a biomarker of atypical network activity in epilepsy, offering a network-specific, whole-brain approach independent of anatomical coordinates. However, its lack of correlation with clinical outcomes underscores the need for further refinement and validation to establish fPSC as a reliable measure of epileptogenic burden.

**HIGHLIGHTS:**

- fPSC analysis using rs-fMRI detects a moderate treatment effect of epilepsy surgery, independent of relative network volume changes.
- fPSC analysis provides a method to quantify whole-brain network dysfunction in epilepsy without the need for a seizure onset zone identification-hypothesis.
- A trend towards direct correlation is observed between fPSC post-operative change and seizure outcomes in epilepsy surgery.

## 1. Introduction

### 1.1 Background

Epilepsy surgery is an effective treatment for drug-resistant epilepsy (DRE)(de Tisi et al, 2011; Téllez-Zenteno et al, 2005; Wiebe et al, 2001). However, outcomes vary greatly because post-operative seizure control depends on the accuracy of the seizure onset zone (SOZ) localization(Englot et al, 2013; Jobst & Cascino, 2015; Juhász & John, 2020). Furthermore, assessing the post-operative seizure reduction is a lengthy process, as success is benchmarked at one-year, or further, from the surgery(Widjaja et al, 2020). Prediction models for epilepsy outcome of epilepsy surgery have been created based on preoperative and postoperative factors associated with good outcomes(Bonilha et al, 2015; Morgan et al, 2017; Negishi et al, 2011; Santos-Santos et al, 2023). While these models provide valuable guidance for surgical decision-making, risk factor-based predictions do not capture the specific impact of the intervention on an individual patient’s condition. Thus, although they are useful for planning, these models are limited in their utility for post-operative follow-up.

Thus, a biomarker is needed to measure the surgery effect on the individual epilepsy network and support the clinical response assessment. Ideally, such a marker would not rely on preexisting SOZ hypothesis or the capture of a seizure, since many are initially seizure free post-operatively, yet go on to have seizure reoccurrence later(Widjaja et al, 2020). This non-ictal, individualized biomarker predicting post-operative seizure outcomes may also inform care through the reduction of the duration of post-operative pharmaceutical therapy with its associated side effects and costs(Cohen-Gadol et al, 2006; de Matos et al, 2023; Eddy et al, 2011).

Success on this front has not been forthcoming. Unfortunately, post-operative single EEG is not predictive of long-term surgical response to guide post-operative pharmaceutical therapy(Menon et al, 2012; Rathore et al, 2011). And though many functional neuroimaging studies in epilepsy have proved useful determining the presence or absence of epilepsy, they are based in groupwise analysis, and little has been translated to individualized clinical follow-up(Jiang et al, 2022; McSweeney et al, 2017; Milton et al, 2022; Nguyen et al, 2021). One promising approach is resting-state functional MRI (rs-fMRI). Rs-fMRI reveals resting-state network (RSN) abnormalities during interictal periods(Hunyadi et al, 2013; Jiang et al, 2022; McSweeney et al, 2017). Abnormal brain networks in DRE patients, as assessed by independent component analysis (ICA), indicate epileptogenic activity and are often referred to as seizure networks (SzNETs). SzNETs spatially correspond to SOZs identified via intracranial stereo EEG in approximately 90% of cases, with a meta-analysis indicating a 71.3% concordance with ground truth (defined as stereo EEG and post-surgical outcomes) for SOZ identification(Boerwinkle et al, 2017; Chakraborty et al, 2020). Moreover, seizure freedom after epilepsy surgery is associated with the reduction of SzNETs and the normalization of the RSN(Boerwinkle et al, 2019).

However, a major limitation of this method is that SzNET identification for clinical implementation currently requires expert visual interpretation of independent components(Banerjee et al, 2023; Hunyadi et al, 2014), as shown in Figure 1. This reliance limits reproducibility and may introduce potential bias (Banerjee et al, 2023; Kamboj et al, 2023). Therefore, there is a critical need for an interictal, individualized, fully automated and reproducible biomarker of pathological epileptic network connectivity that is predictive of post-operative outcomes.

**Figure 1.**
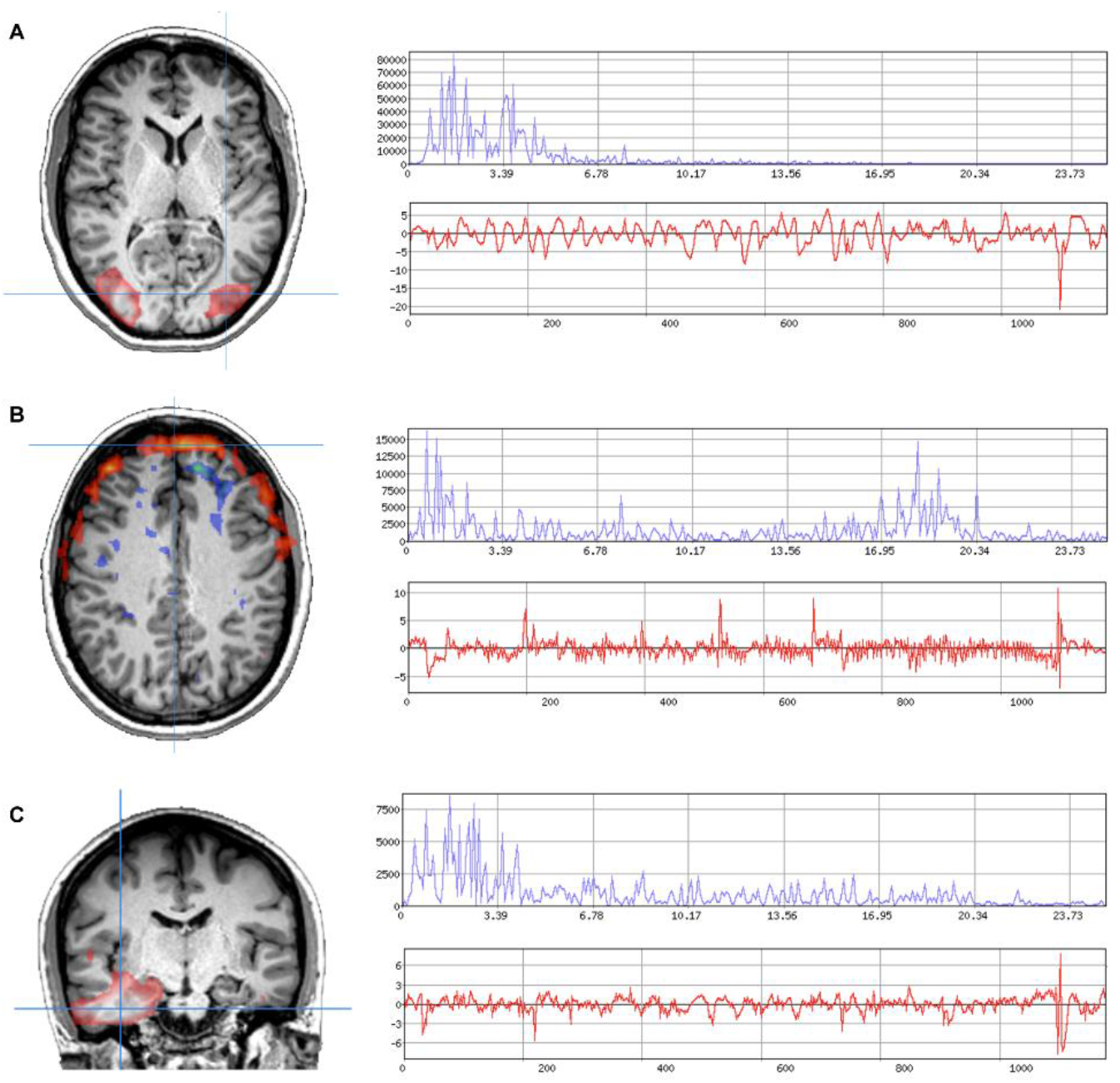
Independent component categories. Each panel displays a unique independent component from the same patient, accompanied by the corresponding power spectrum and BOLD signal time course. The blue plots represent the power spectrum (over Hz/100), while the red plots depict the BOLD signal (over seconds). **Panel A** shows a resting-state network corresponding to a secondary visual network, displaying the characteristic regular BOLD signal. **Panel B** illustrates a noise component associated with motion artifacts, identifiable by its spatial distribution across the brain surface and an irregular BOLD signal. **Panel C** represents a seizure network, characterized by gray matter distribution suggesting neuronal origin and atypical BOLD signal features.

One promising and quantifiable network attribute is the power spectrum amplitude. In SzNETs, power spectrum amplitude is typically higher at elevated frequencies than in RSNs, as observed in the example from Figure 2. Previous studies have shown that voxel-wise power spectrum amplitude features can more effectively distinguish non-lesional epilepsy from healthy controls than structural MRI features and are also useful for classifying independent components identified by ICA(De Martino et al, 2007; McGill et al, 2014). However, power spectrum amplitude has yet to be evaluated post-ICA in relation to epilepsy symptoms or as a measure for quantifying treatment effects.

**Figure 2.**
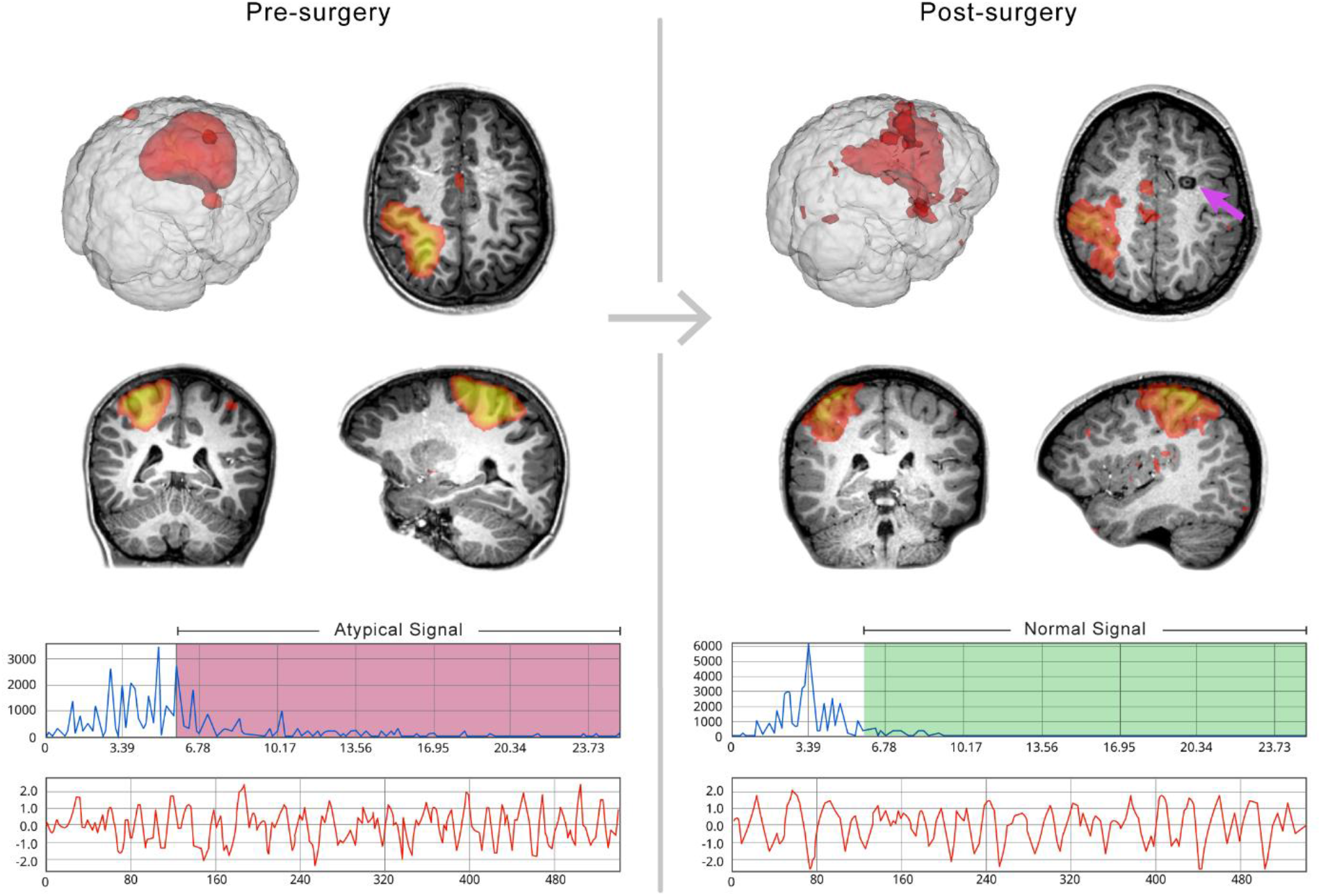
Changes in RSN features following treatment. Pre- and post-surgical changes in resting-state network (RSN) features with similar spatial distributions are shown after laser interstitial thermal therapy applied in a remote location (marked by the magenta arrow). The upper blue plot illustrates the power spectrum, with shaded areas indicating regions used for fractional power spectrum contribution (fPSC) calculations. While the power magnitude varies across networks, a relative decrease in higher frequencies compared to lower frequencies is observed post-surgery. The lower red plots display the BOLD signal, showing a smoother pattern in the post-surgical network.

### 1.2 Objectives

In this retrospective cohort study of DRE patients who underwent epilepsy surgery, we performed individualized quantitative analysis of the RSN’s power spectrum amplitude from rs-fMRI acquired before and after the surgical intervention. For this analysis, we introduced a brain network measure we call fractional power spectrum contribution (fPSC), based on the BOLD spectral properties, designed to characterize the whole brain connectome on the individual level. Our main objectives in this study were to (1) assess the pre-to-post operative changes in fPSC and (2) determine if pre-to-post operative fPSC change correlates with the one-year post-operative seizure reduction. We posited that if the fPSC has potential as a biomarker of epileptogenic network activity, it should improve after effective epilepsy surgery.

## 2. Methods

### 2.1 Data Collection

This study expands upon our previously published work from this retrospective cohort of pediatric epilepsy patients (Boerwinkle et al, 2019). The cohort includes all patients who underwent epilepsy surgery between August 2012 and July 2016 and had both preoperative and postoperative rs-fMRI as part of routine clinical care. All procedures were performed in compliance with relevant laws and institutional guidelines, with approval from the local Institutional Review Board at the time of data collection for the original cohort study(Boerwinkle et al, 2019). The dataset was collected from the medical record by study staff and includes demographics, epilepsy etiology, number of seizure subtypes, epilepsy type, age at first seizure, the date and type of the surgery, seizure frequency at the time of each MRI and seizures improvement at clinical follow-up at least 1-year post-operatively. Scans with unclear current seizure frequency were excluded from the correlation analysis between seizure frequency and fPSC. For subjects with multiple surgical interventions, the date of the last surgical procedure was recorded as the relevant surgery date. Surgical procedures were determined through multidisciplinary epilepsy conferences, which analyzed multiple data modalities to tailor the approach in the best interest of the patient. These procedures included both open and minimally invasive interventions, and were categorized as either non-focal, such as callosotomies and vagal nerve stimulation, or focal, including focal resections, ablations, lobectomies, hemispherectomies, and responsive neurostimulation. There were no cases of thalamic deep brain stimulation.

### 2.2 Rs-fMRI Acquisition

Functional and anatomical MRI of DRE subjects were acquired using a 3T MRI scanner (Ingenuity; Philips Medical Systems, Best, Netherlands) equipped with a 32-channel head coil. Light sedation was used to improve tolerability to the scan and reduce motion artifacts. The pre-and-post operative rs-fMRI sequence parameters were repetition time (TR) = 2000 ms, echo time (TE) = 30 ms, flip angle = 80°, slice thickness = 3.4 mm with no gap, in-plane resolution = 3 x 3 mm, and interleaved acquisition. The total number of volumes acquired was 600, with a total scanning time of 20 minutes. For anatomical reference, a T1-weighted whole-brain sequence was acquired, with TR = 9 ms, TE = 4 ms, flip angle = 8°, slice thickness = 0.9 mm, and in-plane resolution = 0.9 x 0.9 mm.

### 2.3 Preprocessing

Standard preprocessing steps were applied to the rs-fMRI with FMRIB Software library (FSL) tools, including removal of non-brain structures, deletion of the first 5 volumes, motion correction using FSL’s tool MCFLIRT(Jenkinson et al, 2002), inter-leaved slice time correction, high-pass filtering at 100 seconds, and without smoothing. All subjects’ head motion was less than 1 mm in any direction. Individual functional scans were then linearly registered to each subject’s high-resolution T1 scan using linear registration(Jenkinson et al, 2002), and optimization was achieved through boundary-based registration (Greve & Fischl, 2009).

ICA was performed using MELODIC, with the total number of independent components determined by automated Bayesian dimensionality estimation(Beckmann et al, 2005). Manual classification of the components was initially performed to categorize them as RSN, SzNET and noise. The algorithm used for this categorization utilizes features from the spatial and temporal domains and is detailed in prior works(Boerwinkle et al, 2017; Griffanti et al, 2017).

### 2.4 fPSC and volume ratio measurement

The fPSC for each Resting-State Network RSN was quantified using an approach analogous to Zou et al.’s fractional Amplitude of Low Frequency Fluctuations (fALFF) method(Zou et al, 2008). The methodology for calculating fPSC is detailed as follows. fPSC represents the proportion of the power within specific frequencies of interest relative to the total power spectrum. For computational efficiency, the integral of the power spectrum curve between 0.06 and 0.25 Hz was calculated using the ‘*trapz’* function from NumPy(Harris et al, 2020), and divided by the total power spectrum *trapz*- integral of the same RSN, as expressed in the following equation.

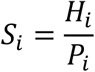

Where *S*_*i*_ represents the fPSC of a network *i, H*_*i*_ is the area under the curve of the power spectrum function between 0.06 and 0.25 Hz, and *P*_*i*_ denotes the total area under the curve for the same RSN’s power spectrum. This calculation yields a single fPSC value per RSN.

Given that ICA with automated Bayesian dimensionality estimation adjusts the number of components (networks and noise) per scan based on signal variability, the total number of RSNs can vary between scans, even within the same subject. To ensure comparability across scans, the fPSC values for all RSNs in a single scan were averaged, providing a representative fPSC value per scan.

Since fPSC measures a proportion, it is independent of the total brain tissue volume. However, many subjects in our cohort experienced brain tissue loss after surgical treatment, raising concerns about potential changes in RSN volumes influencing fPSC values. To address this, we calculated a Normalized Volume Ratio (NVR) to assess whether RSN volumes remained consistent across scans. As with fPSC, variations in the number of RSNs and noise components per scan due to ICA necessitated normalization. The NVR for each network was determined by dividing the number of voxels in the network’s spatial mask by the total number of RSN voxels in the same scan. The average NVR index across all RSNs within each scan was then used to ensure comparability across scans, allowing us to assess RSN volume changes after the treatment.

### 2.5 fPSC Threshold identification

The fPSC focuses on high-frequency components of the power spectrum. To identify the lower boundary of the frequency interval that maximizes the difference between the most abnormal networks (the SzNET) and typical RSNs, we conducted a series of comparisons between expert-derived SzNET-ICs and RSN-ICs using a different dataset of drug-resistant epilepsy patients. These comparisons were made across a range of power spectrum thresholds from 0.01 Hz to 0.2 Hz, in increments of 5 mHz. The threshold of 0.06 Hz was found to provide the greatest median difference in power ratios between SzNET-ICs and RSN-ICs, with a p-value of 0.001. Therefore, the 0.06 Hz threshold was adopted for this study.

### 2.6 Statistical methods

We used descriptive statistics to characterize the study data. Our primary question was whether the average fPSC changes between the pre-treatment scan and the post-treatment scan. We assessed this unconditionally with a paired t-test and controlling for covariates with a linear mixed effects model. Our second question was whether the change in average fPSC is associated with the clinical improvement score. We assessed this with an ANOVA, treating the clinical improvement score as a categorical variable. Finally, the last question was whether the seizure frequency reported at the time of the rs-fMRI is associated with the scan’s average fPSC, while controlling for time (before vs. after). We assessed this with a generalized linear ordinal logistic mixed effects model. Here we treated seizure frequency as an ordinal variable, instead of as a count variable, because the counts were often estimated instead of measured. Both mixed effects models accounted for the non-independence in the data with random intercepts for patients. We first tested models as a whole, and tested constituent variables if the model was significant. Likelihood ratio tests were used for the mixed effects models. An alpha of 0.05 was considered significant.

## 3. Results

A total of 58 patients from the institutional pediatric multidisciplinary epilepsy conferences underwent therapeutic neurosurgical procedures and had both pre- and post-operative rs-fMRI data available. Of these, 56 cases had epilepsy surgery for DRE and were included in the analysis. The remaining two patients received a different type of therapeutic surgery not primary driven for seizure reduction. The demographic details are summarized in Table 1. The median age at surgery was 108.7 months, with a median time from initial symptoms to surgery of 78.6 months. Of the included patients, 80.4% had focal epilepsy, and the most common etiology was hypothalamic hamartoma (37.5%), followed by malformation of cortical development (25%). The median time from the date of surgery to the postoperative scan was 10.7 months. 80.4% of the patients had more than 50% improvement in seizure control and 64.3% were seizure free at clinical follow up time. The outcome variables are described, both overall and stratified by time point, in Table 2.

**Table 1.** Demographics. Table 1. Categorical variables (e.g., sex) are represented by counts with percentages in parentheses. Continuous variables are represented by medians with the interquartile range in square brackets. HH, Hypothalamic hamartoma, TBI, Traumatic Brain Injury. * Focal surgeries exclude vagus nerve stimulation and callosotomies

| Variable | Overall |
| --- | --- |
| **n** | 56 |
| **Age onset** | 12.0 [7.0, 20.5] |
| **Age presurgical scan** | 108.0 [55.8, 168.6] |
| **Lag from onset to surgery** | 78.6 [47.8, 110.8] |
| **Age at surgery** | 109.7 [60.9, 174.8] |
| **Surgery to post scan (days)** | 326.0 [96.8, 425.8] |
| **Clinical improvement** |  |
| None | 3 (5.4) |
| <50% | 8 (14.3) |
| >50% | 9 (16.1) |
| 100% | 36 (64.3) |
| **Handedness** |  |
| Right | 28 (50.0) |
| Left | 22 (39.3) |
| Undefined | 6 (10.7) |
| **Sex** |  |
| Female | 25 (44.6) |
| **No. of seizure types** |  |
| 1 type | 22 (39.3) |
| Multiple | 33 (58.9) |
| Unknown | 1 (1.8) |
| **Epilepsy etiology** |  |
| unidentified | 1 (1.8) |
| HH | 21 (37.5) |
| Neoplastic by histopathological exam | 8 (14.3) |
| Migration defect | 14 (25.0) |
| ischemic | 3 (5.4) |
| hemorrhagic | 1 (1.8) |
| TBI | 1 (1.8) |
| genetic | 3 (5.4) |
| Post-infection | 2 (3.6) |
| encephalocele | 1 (1.8) |
| autoimmune | 1 (1.8) |
| **Epilepsy type** |  |
| Focal | 45 (80.4) |
| Generalized | 2 (3.6) |
| Both | 9 (16.1) |
| **Type of surgery (focal\*)** |  |
| Yes | 53 (94.6) |
| No | 3 (5.4) |

**Table 2.** Key Outcome Variables. Table 2. Characteristics of outcome variables at the individual measurement level. Continuous variables are presented as means (SD) for normally distributed data or as medians (interquartile range) for non-normal data (seizure frequency). p-values for load values are derived from paired t-tests, while the p-value for seizure frequency is based on the Wilcoxon signed-rank test. fPSC, fractional power spectrum contribution; NVR, normalized volume ratio.

| Variable | Overall | Time |  | p |
| --- | --- | --- | --- | --- |
|  |  | Pre | Post |  |
| n | 112 | 56 | 56 |  |
| Mean fPSC | 0.3 (0.1) | 0.34 (0.10) | 0.29 (0.09) | 0.005 |
| Mean NVR | 0.0 (0.0) | 0.0 (0.0) | 0.04 (0.01) | 0.45 |
| Seizure frequency | 4.5 [0.0, 30.0] | 20.0 [4.0, 35.0] | 0.0 [0.0, 1.0] | <0.0001 |

There was no statistically significant difference in the mean NVR of the RSN before and after treatment (mean difference=0.003, p=0.45, Hedges’ g=0.08, 95% CI= [-0.18, 0.35]). The post-treatment average fPSC values differed from the pre-treatment values (t=3.0, df=55, p=0.005, Hedges’ g = 0.40, 95% CI= [0.12, 0.68]) when tested unconditionally, as shown in Figure 3. When controlling for covariates (the lag between onset and epilepsy surgery, the epilepsy type, and sex), the model was significant (χ2=12.2, df=5, p=0.033). The average fPSC differed by time (after vs. before: χ2=8.4, df=1, p=0.004), and the associations with the other covariates were not found significant (all p-values >0.10). We considered using the surgery type as a covariate to be included in the model, but it was highly correlated to the epilepsy type and therefore not included to avoid collinearity.

**Figure 3.**
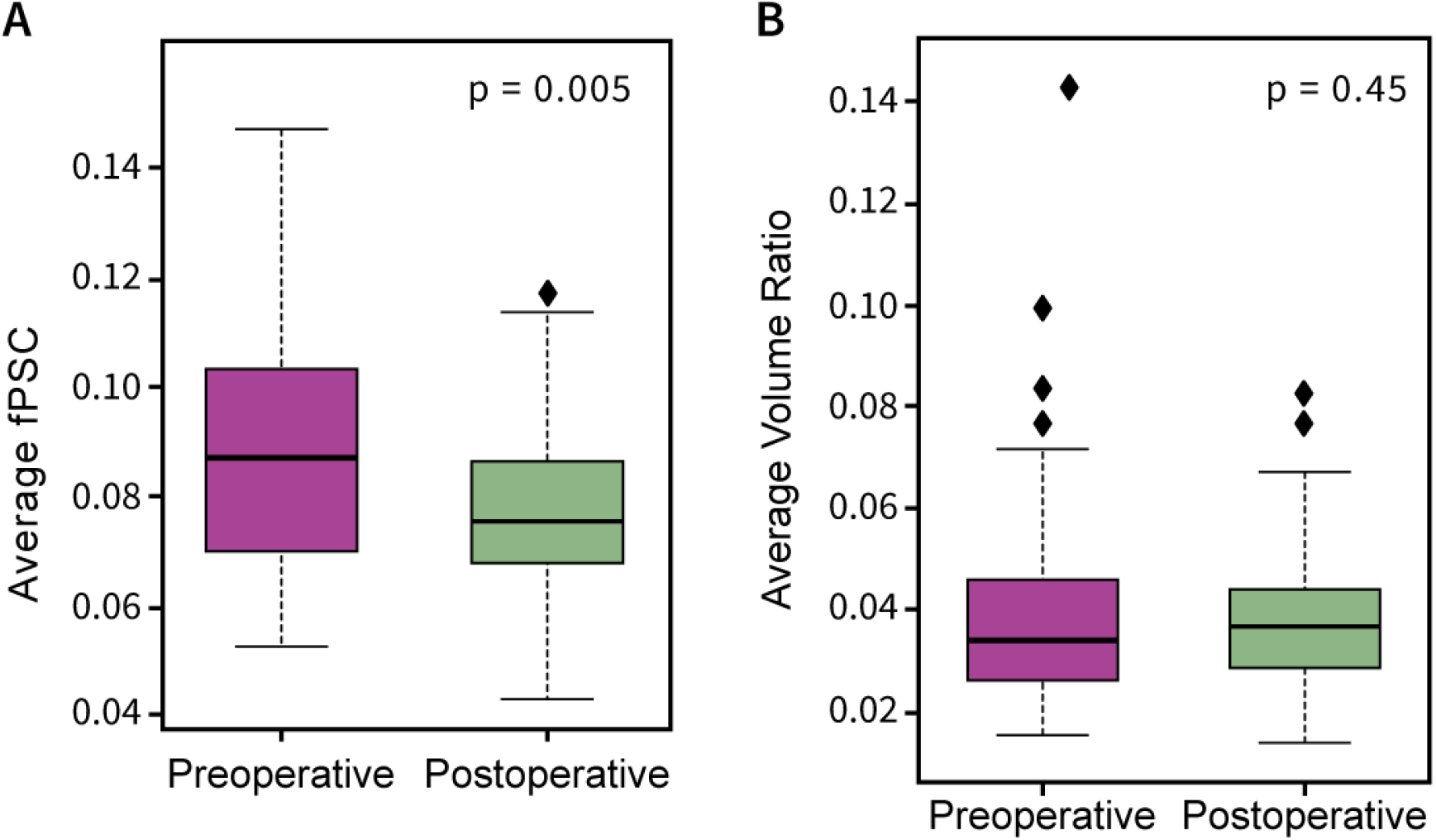
Postoperative changes in RSN outcome variables. Boxplots display the averaged pre- and postoperative values for key outcome variables, with each data point representing the mean resting state networks (RSN) values per patient. **Panel A** illustrates changes in fractional power spectrum contribution (fPSC), while **Panel B** shows changes in normalized volume ratio (NVR). Whiskers extend to the most distant point within 1.5 times the interquartile range, with more extreme values plotted individually. *P*-values were calculated using paired *t*-tests.

There was insufficient evidence to determine an association between changes in average fPSC and the clinical improvement score (F=1.8, dfn=3, dfd=52, p=0.16). However, a trend was noted between the postoperative fPSC change and clinical outcome, as shown in Figure 4. The model for the association between the average fPSC and the seizure frequency was significant (χ2=56.0, df=2, p<0.0001), however there was insufficient evidence to determine an association between the fPSC and the frequency (z=-0.7, p=0.49).

**Figure 4.**
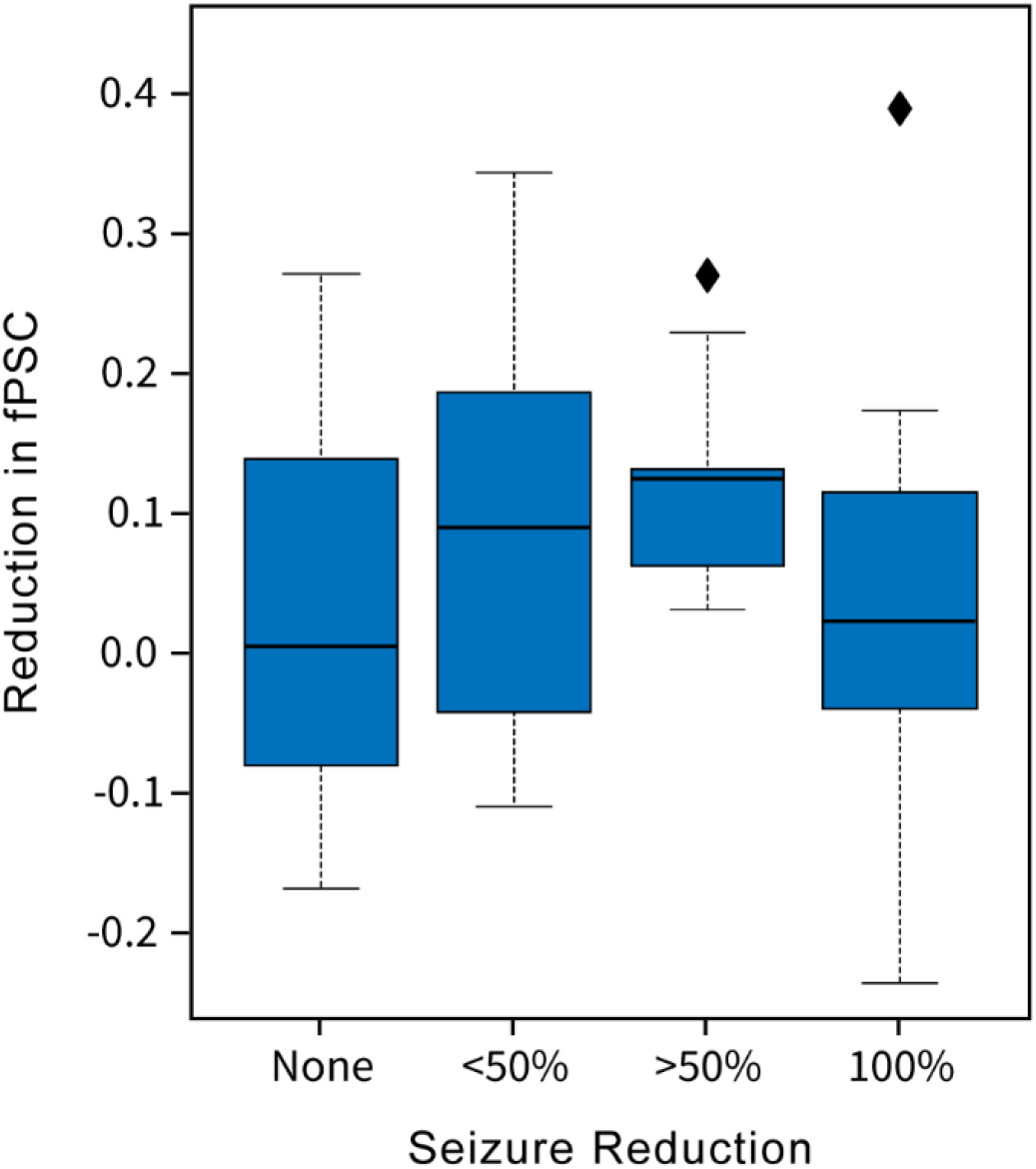
Seizure outcome in relation to connectivity changes. Boxplots illustrate the difference in mean fractional power spectrum contribution (fPSC) between pre- and post-surgical scans, stratified by seizure reduction percentage at a minimum of 1-year follow-up. Whiskers extend to the most distant point within 1.5 times the interquartile range, with more extreme values plotted individually.

## 4. Discussion

The current cohort presents a heterogeneous group of epilepsy surgery patients with overall good surgical outcomes. The significant reduction in fPSC observed after surgical treatment suggests an association with the effectiveness of these interventions. However, this retrospective study did not demonstrate a correlation between fPSC changes and seizures improvement, which may be due to variability in the timing of postoperative imaging and clinical follow-up assessments, among other factors. Interestingly, spectral features of the BOLD time course, rather than volumetric properties of the RSN, were associated with treatment effects. This is noteworthy because treatments primarily involving ablative techniques were expected to affect cluster volumes, either due to brain tissue loss or reduced capacity to detect true signals from the data. However, the power spectrum variable proved to be more informative than the volumetric variable.

Numerous studies have highlighted the biomarker potential of the frequency and amplitude of the power spectrum in the BOLD signal for epilepsy, such as those utilizing the fALFF(Chen et al, 2017; Gupta et al, 2017; Li et al, 2023; Wang et al, 2018). fALFF, derived from the power spectrum ratio of a specific frequency band over the power spectrum of the entire observed frequency range(Zuo et al, 2010), has recently been used as a marker of pathological states and to assess functional connectivity(McGill et al, 2014; Zhang et al, 2023). It provides insights even in the absence of anatomical imaging findings, as seen in cases of temporal lobe epilepsy(Song et al, 2024), and shows promise in distinguishing different zones of epileptogenic networks, as demonstrated in the pilot study by Sathe et al(Sathe et al, 2023). Additionally, fALFF has proven effective in identifying treatment effects across various interventions, including pharmacological treatment(Qiao & Niu, 2017; Yan et al, 2020) and vagal nerve stimulation(Zhu et al, 2020).

While the power band analyzed was defined by SzNET features, it is unclear why the high-frequency power band was particularly informative of the therapeutic effect of epilepsy surgery. Lower power bands between 0.01 and 0.1 Hz have been identified to reflect neuronal signals(Balduzzi et al, 2008; Buzsaki & Draguhn, 2004; Vanhatalo et al, 2004), but in pathological conditions like epilepsy, relevant information may exist outside these low-frequency bands, although the underlying reasons remain unknown.

### 4.1 Comparability challenges

It is frequent for fALFF studies to compare parametric maps generated voxel-wise using healthy controls, aiming to identify brain regions with specific activation patterns or detect changes following interventions. However, these innovative approaches face challenges due to the inherent heterogeneity of epilepsy and its variable epileptogenic networks across patients. Even clinically similar epilepsies can exhibit different anatomical distributions of their seizure onset zones and symptomatogenic zones(Tufenkjian & Lüders, 2012; Turek & Skjei, 2022), making it difficult to find statistically significant results and generalize them to other epilepsy patients based on coordinate systems. These replicability and comparability issues are prevalent in the rs-fMRI field, as highlighted by the ENIGMA Brain Injury working group, which calls for more metrics that can be applied at the individual level and compared across subjects(Caeyenberghs et al, 2024).

Nevertheless, the analysis of power bands has been applied to established RSNs and has shown some utility in classifying these networks and assessing treatment effects(De Martino et al, 2007; Obst et al, 2022). Therefore, the spectral features of functional networks represent a potential source for identifying biomarkers of pathological activity. Our methods aim to leverage dimensionality reduction from ICA to select relevant signals using the spatial domain (selecting the RSNs) and subsequently use a quantitative pathological feature from the signal to derive a network-specific index normalized for each network, without relying on their anatomical coordinates for comparisons. This approach enables further dimensionality reduction, allowing us to work with a standardized metric suitable for individual-level analyses while still conducting a whole-brain connectome-level analysis. This whole-brain approach is crucial when dealing with epileptogenic activity because seizures are highly heterogeneous and not always localized to the same brain regions. Therefore, targeting a specific subnetwork associated with the symptom, as is often done in rs-fMRI studies of other diseases, may not be feasible.

While we have attempted to address the comparability challenges associated with the spatial location of signals, other sources of heterogeneity will require additional strategies. For instance, disease heterogeneity may explain the lack of correlation between fPSC and seizure frequency, despite both variables showing significant reductions following treatment. The spectral properties of functional connectivity with epileptogenic potential may vary depending on factors such as epilepsy etiology or the number of seizure types, given that different forms of epilepsy are associated with distinct EEG abnormalities(Westmoreland, 1998). Since BOLD signal oscillations align with local field potentials(Logothetis et al, 2001), these variations in neuronal activity across epilepsy etiologies may result in differing BOLD oscillation patterns. Unfortunately, the sample size of our study did not allow for corrections based on those confounding variables, despite their collection. Additionally, our cohort faces selection bias, as more complex cases or etiologies associated with specialized clinical programs, such as hypothalamic hamartoma, may be more likely to undergo post-surgical rs-fMRI follow-up imaging.

### 4.2 Automation relevance

To effectively test this and similar methods with larger datasets, including those from different institutions, automating the process is crucial. Identifying SzNETs has been a challenging step for using automated pipelines due to the need for expert assessment. One additional advantage of our approach is that SzNET identification was not required beyond setting the threshold for the power band. This is particularly beneficial since current machine learning algorithms are more effective at identifying RSNs than SzNETs(Banerjee et al, 2023; Hunyadi et al, 2014; Kamboj et al, 2023; Nozais et al, 2021).

The lack of consensus on denoising strategies further highlights the need for network identification processes that can be fully automated. Testing multiple denoising methods could yield additional insights, and some of these steps may need to be applied before final network identification, potentially requiring repetition of the process. While we acknowledge that incorporating additional denoising steps might have enhanced our signal quality and results, they were not included in this study to maintain consistency with the validated methods previously established by our lab for SzNET identification.

In conclusion, our study highlights the potential of spectral features derived from functional networks as biomarkers of pathological activity in epilepsy. While our approach benefits from the dimensionality reduction provided by ICA and focuses on spectral characteristics rather than anatomical coordinates, the challenges posed by the inherent heterogeneity of epilepsy remain significant. Although our findings suggest that fPSC reduction after surgical intervention correlates with treatment efficacy, the lack of association with clinical outcomes underscores the need for further refinement of these methods. To fully realize the potential of power spectrum-based biomarkers, future research should prioritize larger, more homogenous cohorts, automation of analytic processes, and the integration of additional denoising steps. These efforts will be critical for developing reliable, individualized metrics that can be applied across diverse patient populations and improve our understanding of treatment effects in epilepsy.

### Glossary

#### Blood Oxygenation Level Dependent (BOLD)

A signal useful to extrapolate brain activity detected through MRI that reflects changes in blood oxygen levels, it is correlated with local field potential in the brain.

#### Independent Component Analysis (ICA)

A statistical method used to separate mixed signals into independent sources. In neuroimaging, ICA is commonly used to isolate brain networks and separate them from noise or artifacts in fMRI data.

#### Independent Component (IC)

An output of ICA, representing a spatially distinct brain network or artifact. Each IC consists of a unique spatial map and time course, used to identify functionally related brain regions and isolate noise sources in neuroimaging data.

#### Power Spectrum

A representation of the distribution of signal power across frequency bands. In fMRI, power spectrum analysis quantifies the contribution of different frequency ranges to the brain’s BOLD signal fluctuations.

#### Seizure Onset Zone (SOZ)

The brain region where epileptic seizures originate, identified to target surgical interventions in epilepsy.

## Data Availability

The data will be made available upon reasonable request.

## Declaration of generative AI and AI-assisted technologies in the writing process

During the preparation of this work the author(s) used Copilot and ChatGPT in order to proofread and improve the clarity and readability of the writing. After using this tools, the authors reviewed and edited the content as needed and take full responsibility for the content of the publication.

